# FooDOxS: A Database of Oxidized Sterols Content in Foods

**DOI:** 10.1101/2023.11.15.23298592

**Authors:** Ilce Gabriela Medina-Meza, Yashasvi Vaidya, Carlo Barnaba

**Author notes:** Corresponding Authors Medina-Meza Ilce Gabriela, PhD. Department of Biosystems and Agricultural Engineering, Michigan State University 469 Wilson Rd.| Room 302C East Lansing, MI Carlo Barnaba, Ph.D. Department of Pharmaceutical Chemistry, University of Kansas 2030 Becker Dr.| Room 320D Lawrence, KS. Authors I.G.M.M. and Y.V. contribute equally to this paper. Current affiliation: *Department of Molecular, Cellular, and Developmental Biology, University of Michigan, Ann Arbor, USA.

## Abstract

This research addresses the knowledge gap regarding dietary oxidized lipids (DOxS) in foods classified under the NOVA and WWEIA systems. We present the FooDOxS database, a comprehensive compilation of DOxS content in over 1,680 food items from 120 publications across 25 countries, augmented by internal lab data. Our analysis discerns DOxS exposure in diverse diets, differentiating between plant-based and animal-based sources. Notably, we evaluate the efficacy of NOVA and WWEIA classifications in capturing DOxS variations across food categories. Our findings provide insights into the strengths and limitations of these systems, enhancing their utility for assessing dietary components. This research contributes to the understanding of DOxS in food processing and suggests refinements for classification systems, holding promise for improved food safety and public health assessments.

## 1. INTRODUCTION

Consumer awareness of food ingredients is a fundamental pillar in the quest for healthier eating habits and maintaining overall well-being. Consumers conceptualize ‘healthy’ foods based on food characteristics, specifically food groups, nutrients, production or processing, or lack of specific ingredients^1^. This awareness has the potential for individuals to make informed decisions, navigate dietary preferences, and take control of their health^2,3^. Nutrient databases serve as an invaluable tool in enhancing both consumer awareness and nutritional interventions. Such databases aid in dietary analysis for individuals and groups, intervention material development, and menu planning for studies^4^. Above all, the accuracy of food composition information within the database is paramount^5^. Comprehensive nutrient databases, mainly focusing on macronutrients, are relatively widespread^6^. However, databases for essential micronutrients, particularly those with established nutritional and biological significance, remain scarce^7^. In the context of food safety, public health, and regulatory oversight, establishing comprehensive food contaminant databases is of paramount importance. These databases play a pivotal role in monitoring and mitigating potential risks associated with contaminants entering the food supply chain^6^. For example, databases specifically addressing compounds like acrylamide have driven the development and implementation of strategies aimed at controlling its formation and reducing its presence in a variety of food products^8^. Our laboratory has extensively studied dietary oxysterols (DOxS), a group of molecules derived from their parent compound -cholesterol or phytosterol-, with an additional hydroxyl, ketone, or epoxy group^9–11^. Oxysterols can be enzymatically produced in the body by the cytochrome P450 family, serving as intermediaries and activators of cell-signaling pathways^9^. Alternatively, non-enzymatic production can occur due to oxidative stress^9^. DOxS are known to exert pro-inflammatory, pro-oxidant, pro-fibrogenic, and pro-apoptotic toxic effects, leading to chronic diseases like atherosclerosis, hypertension, Huntington’s disease, Parkinson’s disease, multiple sclerosis, Alzheimer’s disease, and some cancers, among others^9,12–14^. DOxS derived from phytosterols are suspected to have a similar effect due to the structural similarity of phytosterol and cholesterol^15–17^.

There is a consensus that the formation of DOxS is induced by processing, primarily due to temperature and the presence of radical species, which undeniably influence their formation kinetics^10^. Light exposure, radiation, excessive storage, and other agents that lower the activation energy of the reaction unintentionally contribute to DOxS generation. Consequently, the prevalence of DOxS in processed foods has become a significant concern^9,18,19^. The high levels of processing facilitate DOxS formation, thereby increasing consumer exposure to them. For instance, our laboratory has conducted assessments of DOxS dietary exposure in infants fed with various milk formulas, revealing that exposure levels are contingent on the extent of food processing applied to the formula. Thus, the relationship between processing and DOxS formation underscores the need for improved control measures and awareness in food production to mitigate potential health risks.

Little is known about the presence of dietary oxidized lipids (DOxS) in foods categorized under the NOVA food classification system, which is a widely accepted framework used for classifying foods based on the nature and extent of their processing^20^. This classification method, introduced in 2009 by Monteiro’s group, has become a crucial tool for investigating the health impacts of food processing on various chronic diseases. While NOVA has revolutionized our perspective on food by emphasizing the level of processing involved, it has also faced critiques from various quarters, particularly in its characterization of ultra-processed foods (UPFs)^21,22^.

This research aims to address the existing gaps in knowledge by constructing FooDOxS, a comprehensive database detailing the content of dietary oxidized lipids (DOxS) in more than 1,680 food items, each meticulously classified under the NOVA and WWEIA systems. Drawing on data from 120 publications across 25 countries and supplementing it with information from the lab’s internal database, the resulting FooDOxS database provides a nuanced understanding of DOxS exposure in diverse diets, allowing for discernment based on food sources—whether plant-based or animal-based. This holistic approach not only contributes to our understanding of DOxS in relation to food processing but also facilitates a critical examination of NOVA’s and WWEIA’s effectiveness in capturing variations in DOxS content across different food categories. By shedding light on the strengths and limitations of these classification systems, the research underscores the potential for refining and optimizing them to better serve as tools for assessing dietary components and their implications for public health.

## 2. MATERIALS AND METHODS

### 2.1 Literature Search and Data Collection

DOxS data was obtained from an extensive literature search using the terms “cholesterol oxidation products”, “COPs”, “oxysterols”, “phytosterols oxidation products”, and “dietary oxysterols”. Data search and entry were conducted from 2018 to 2022, with studies performed between 1984-2022. The database also includes data from our group previously published^23,24^. In the FooDOxS database, we included studies that specifically analyzed **a)** foods from animal and plant sources; **b)** commercially available foods, **c)** experimentally prepared samples. We took into consideration studies published in 3 languages including English, French, and German, performed in 27 countries including Australia, Austria, Belgium, Brazil, China, Denmark, Finland, France, Germany, Hungary, India, Ireland, Italy, Japan, Jordan, Mexico, Poland, South Korea, Spain, Sweden, The Netherlands, Switzerland, Taiwan, United Kingdom, and the United States. Excluded studies from this database consisted of **i)** literature reviews; **ii)** those reporting DOxS concentration per gram of cholesterol without providing the amount of cholesterol used for analysis; **iii)** papers reporting concentrations as a graph only (not quantitative amounts); **iv)** papers reporting DOxS concentrations as a range.

All values were converted into uniform units to facilitate comparison: µg/g of sample for DOxS and mg/g of sample for phytosterols. These units were the most encountered in the analyzed studies, but some studies also reported DOxS concentrations per gram of lipids or per gram of cholesterol. In this case, the reported quantity of lipids used, or the concentration of cholesterol measured, was used for calculation respectively.

### 2.2 Application of NOVA Criterion

The FooDOxS database follows the NOVA classification as shown in **Figure 1**. The NOVA classification categorizes foods into four groups: 1) unprocessed or minimally processed foods, 2) processed culinary ingredients, 3) processed foods, and 4) ultra-processed foods.^25,26^ Compared to the original NOVA decision flowchart^25,27^ our criterion **a)** highlights individual unit operations as they contribute to processing level and **b)** distinguishes between group 2 additives and industrial additives in the classification of processed vs ultra-processed foods. One of the key questions pertains to the categorization of food as ’convenience foods.’ This category encompasses any food item in which, as per Scholliers, ’the degree of culinary preparation has been taken to an advanced stage, and these items are typically purchased as labor-saving alternatives to less highly processed products’^28^. This initial step facilitates the distinction between homemade and industrial foods.

**Figure 1.**
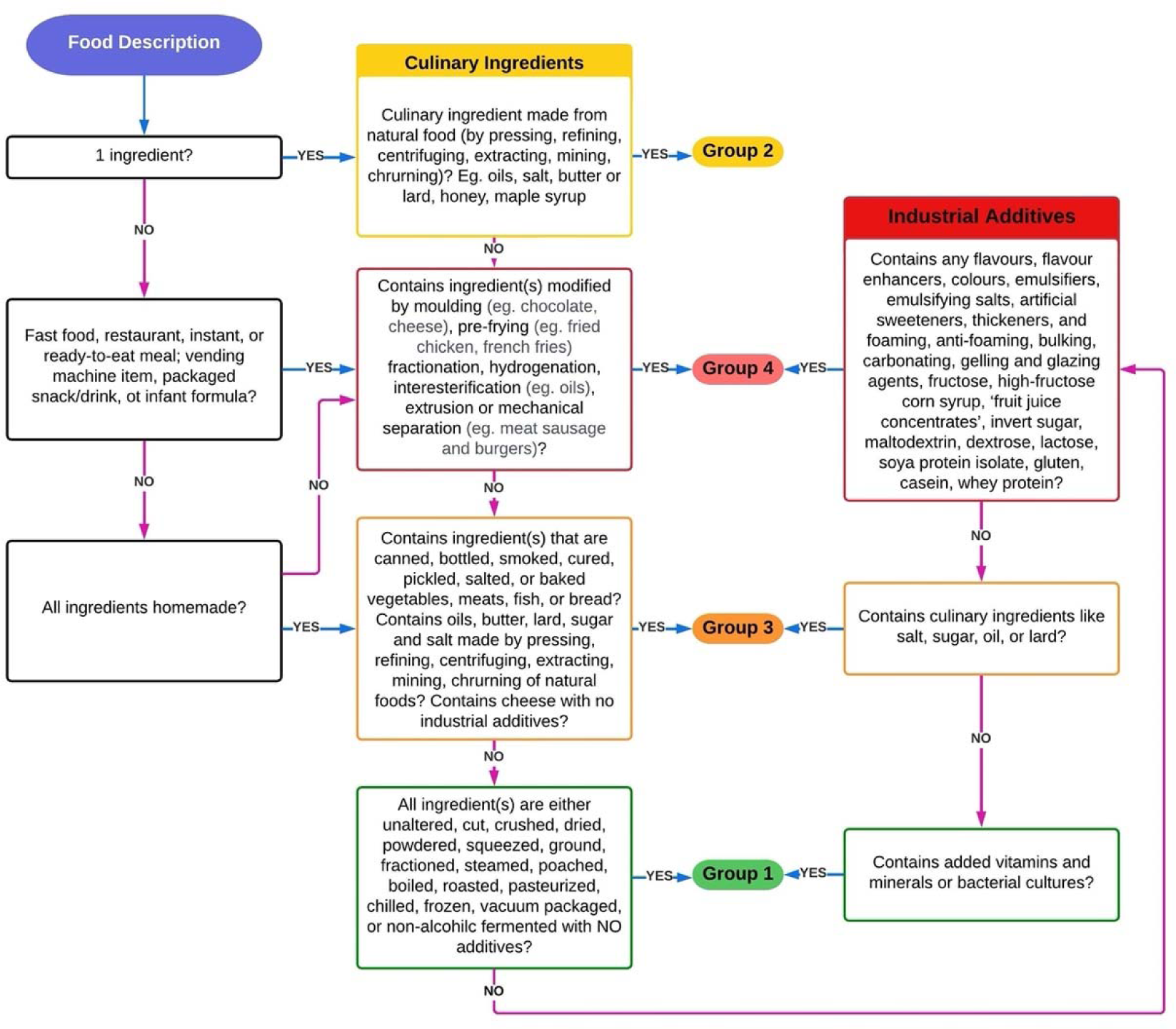
NOVA classification. Flowchart for classifying food items according to the NOVA classification criteria

### 2.3 Assignment of food categories according to What We Eat In America (WWEIA)

We also used The What We Eat in America (WWEIA) food categorization scheme as referenced to assign a category to each food item **(Supplement 2).** This scheme is designed to be applied to foods consumed in the American diet^29^. Since the FooDOxS database contains foods from all around the world, the specific WWEIA codes did not accommodate every food item. However, the main and sub-categories serve as an adequate general search criterion for the user and were thus implemented. All items are categorized by source (animal- or plant-based),

WWEIA food group, and WWEIA main food category. WWEIA codes pertaining to sub-food categories are assigned to each item.

### 2.4 Navigating the FooDOxS database

The FooDOxS database is organized as a spreadsheet in which row represents an individual food item, and column representing either a group variable or dependent variable (i.e., DOxS specie). Specifically:

- *Column A*: a detailed description of each item including processing time and/or temperature, storage time and/or temperature, and any other relevant details reported by the original study.
- *Column B*: the NOVA classification of each food item.
- *Column C*: Serving size if reported in the original study.
- *Column D*: WWEIA food code corresponding to sub-food categories.
- *Column E*: In text-citation of references.
- *Column F*: Additional notes on the food item if present

From *Column G*, DOxS concentration is reported as mean and standard deviation in separate columns. Users can filter the data by a selected column to view food items assigned to a specific Food Group, Main Food Category, NOVA group, or reference. The database has been deposited in the FHEL GitHub page: https://github.com/FHELMSU/FooDOxS.

### 2.5 Statistical analyses

For comparing DOxS content across group categories, a Kruskal-Wallis ANOVA was performed, followed by a Dunn test to estimate mean rank differences, with a significance level set at p = 0.05. Statistical tests were performed in OriginPro v.2023 (OriginLab).

## 3. RESULTS

### 3.1 The FooDOxS database

The aim of this study is to establish a database of DOxS contents in food items classified under the NOVA food classification system. These data were collected from various sources, combining the results of these studies into a single repository, the FooDOxS database. This database contains DOxS data from n = 120 studies dating from 1984 to 2022. Food items were categorized according to the NOVA flowchart (**Fig. 1**), resulting in a total of 1,676 items. These items were distributed as follows: Group 1, 31.1%; Group 2, 19.3%; Group 3, 15.0%; Group 4, 34.6%. Seventy-three percent were of animal origin, while the remaining 27% originated from plants (**Fig. 2A**). We further classified food items using the What We Eat in America classification chart (**Fig. 2B**)^30^. The main categories included protein-based foods (35.1%), fats and oils (26.4%), and dairy (17.2%), reflecting the extent of DOxS research in these categories. A total of 81 different compounds were reported, including parental sterols (i.e., cholesterol, β- sitosterol, campesterol) and DOxS (**Table 1)**. In early studies on cholesterol oxidation, it was often challenging to distinguish between oxysterol isomers. For example, this challenge was evident in cases such as 7α-OH/7β-OH and 5α,6α-epoxy/5β,6β-epoxy, which were frequently reported as mixtures of isomers. To account for this discrepancy, we provided an additional entry that sums those isomers. It is worth noting that we obtained DOxS content data for various items, including 30 infant formulas and over 60 ultra-processed foods, in our laboratory, and these results were previously published^23,24^.

**Figure 2.**
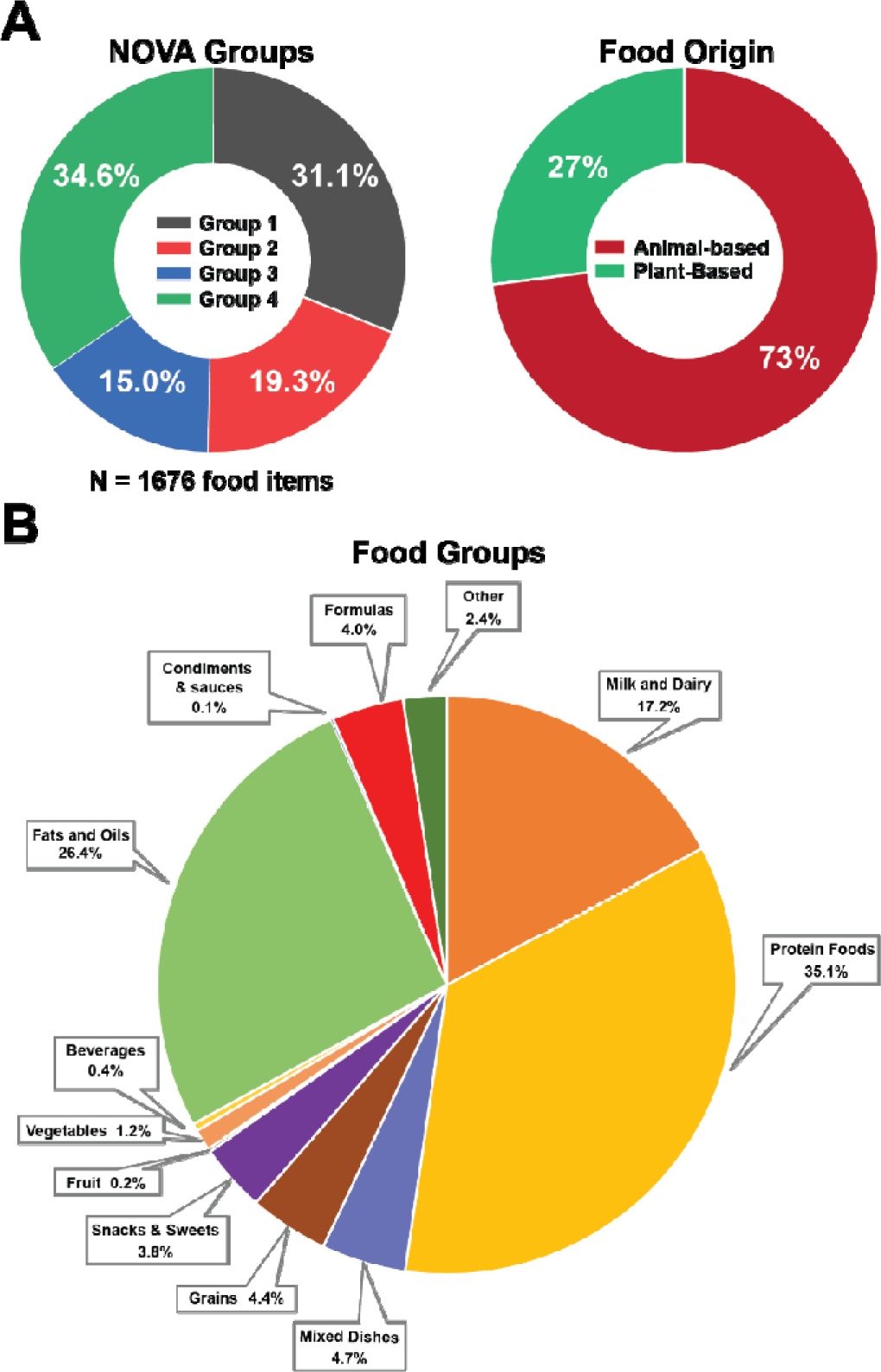
Insights into the FooDOxS database. **(A)** Percentage distribution of food items according to NOVA classification and source (animal vs plant). **(B)** Percentage distribution of food items according to food source.

**Table 1.**
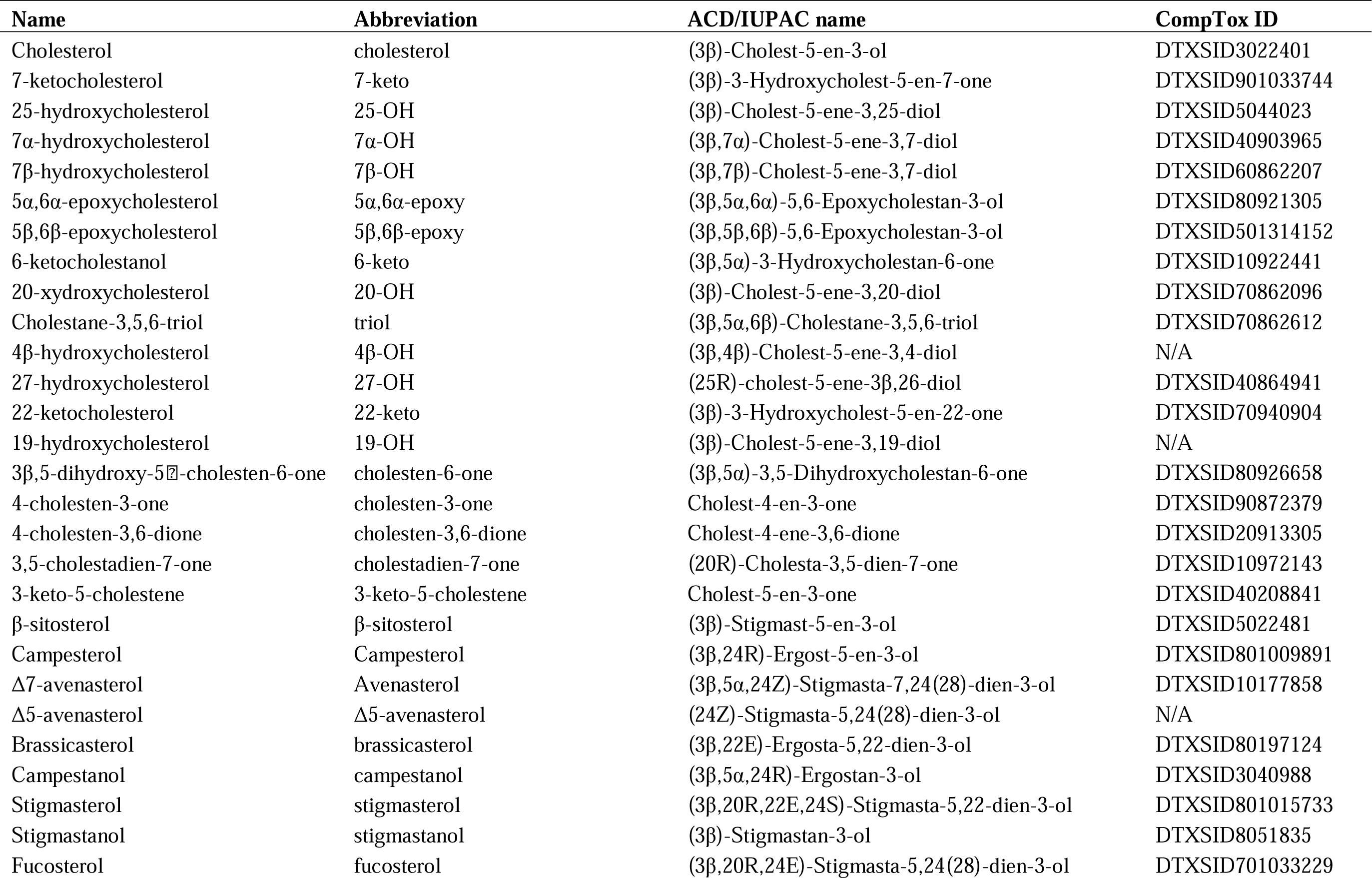
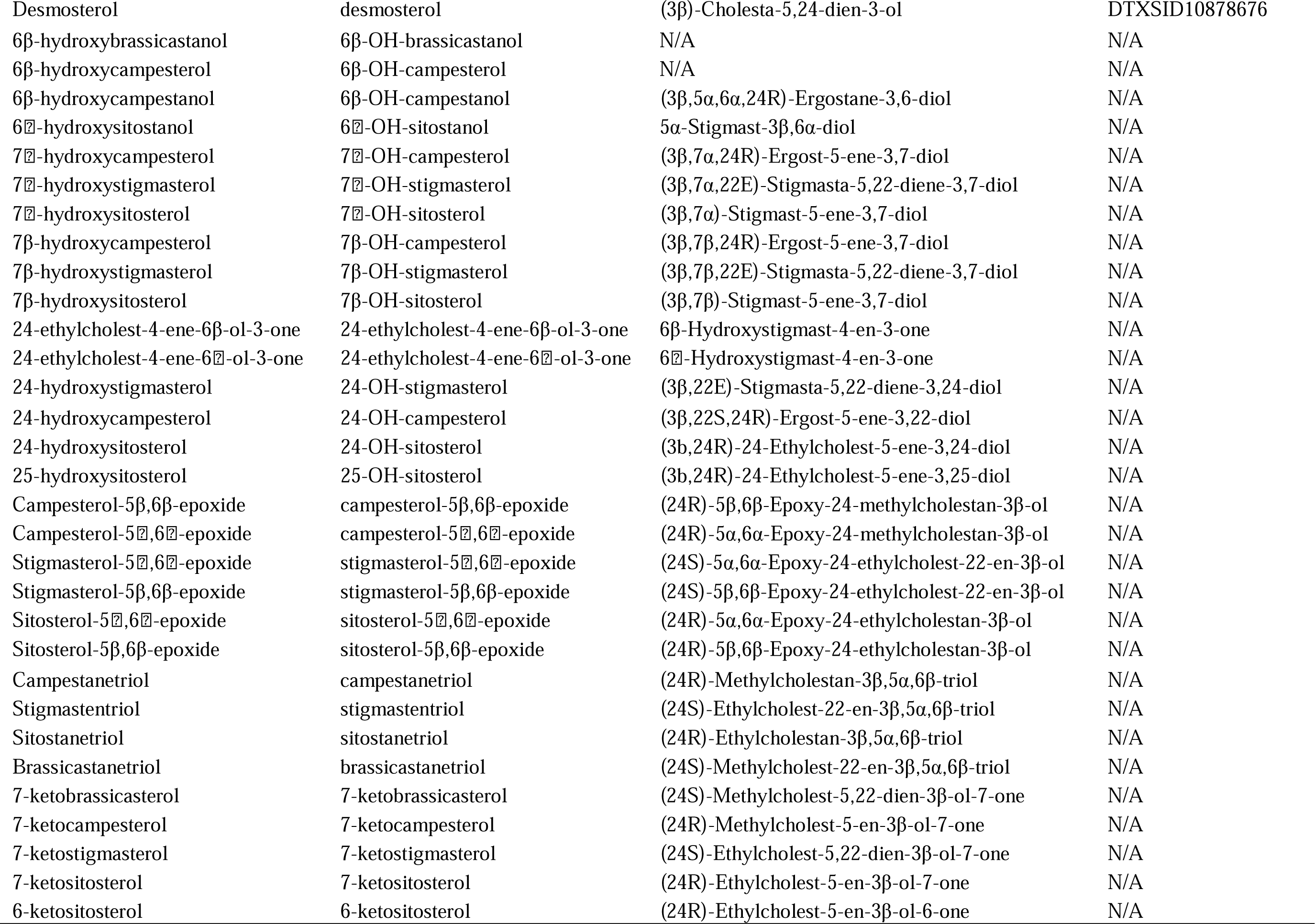
List of compounds included in the FooDOxS database, including common name, abbreviations used in the database, IUPAC systematic name and EPA CompTox ID. ^31^**. N/A = CompTox entry not available.**

### 3.2 DOxS content in animal and plant-based foods under NOVA classification

The creation of an expanded food database opens avenues for a more comprehensive estimation of compound levels across various food categories. Databases become particularly compelling for molecules with recognized bioactive properties, enabling in-depth toxicological and epidemiological studies. These studies include critical aspects like dietary exposure and risk assessment. We examined DOxS concentrations in both animal-based and plant-based foods (**Fig. 3**), stratified according to the NOVA flowchart (**Fig. 3A**) Our initial analysis involved the concentration of all DOxS, achieved either by aggregating all individual oxidative derivatives reported within a food item or by utilizing the total reported DOxS, within NOVA classification. In animal-based foods (**Fig. 3B**), DOxS concentrations varied significantly, ranging from less than 1 ng/g to over 4 mg/g. Surprisingly, when employing a non-parametric ANOVA to compare food categories based on NOVA classifications, group 4 (ultra-processed foods) exhibited lower DOxS amounts compared to all other groups (*p* < 0.001). Next, our attention turned to the 7-OH isomers, 7-keto, and 5,6-epoxy isomers. Due to limitations in some older reports that could not distinguish between these isomers, we opted to combine them. Intriguingly, ultra-processed foods (group 4) exhibited significantly lower amounts of 7-OH and 7-keto compared to minimally processed foods (group 1) and processed culinary products (group 2) (**Fig. 3C,D**). In the case of 5,6-epoxy, the notable distinction was between groups 4 and 1, with the former being richer in these compounds (*p* < 0.05) (**Fig. 3E**). We then focused on plant-based foods, constituting approximately a quarter of the total food items in the FooDOxS database (**Fig. 2A**). Our initial exploration involved mapping phytosterols’ content, the parent compounds of DOxS in plants (**Fig. 3F**). Notably, both processed culinary ingredients (group 2) and ultra-processed foods (group 4) exhibited higher concentrations of phytosterols compared to minimally processed foods. Unfortunately, insufficient sampling for group 3 hindered a fair statistical comparison among the NOVA categories. Similarly, the scarcity of data in the literature regarding plant-based DOxS, commonly denominated as ’phytosterols oxidation products’ or ’POPs,’ limits a comprehensive comparison. For the available data in groups 2 and 4, where the majority of DOxS data exist, ultra-processed foods exhibited higher DOxS content compared to processed culinary ingredients (**Fig. 3F**, right panel). This suggests that processing has a more pronounced impact on DOxS accumulation in plant-based foods than in animal-based ones. However, the lack of studies for plant-based processed foods (NOVA group 3) emphasizes the need for further research to better understand their DOxS accumulation.

**Figure 3.**
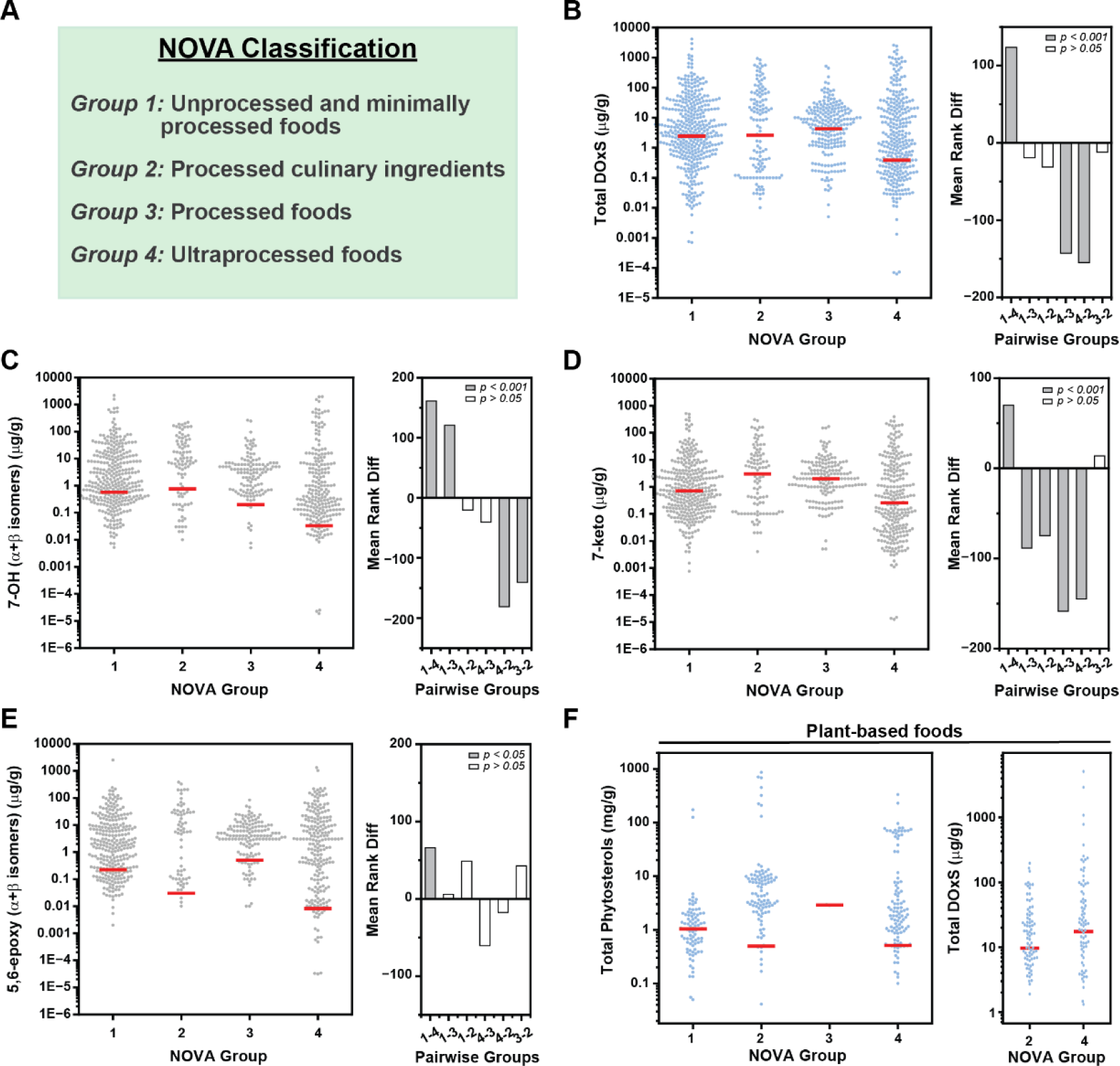
**DOxS abundance in FooDOxS database according to NOVA classification**. **(A)** NOVA groups. **(B)** Left: dots plot showing total DOxS content in animal-based foods according to NOVA classification. Red line represent median. Right: Kruskal-Wallis mean rank differences between groups at *p* = 0.05 significance level. **(C)** Left: dots plot showing 7-OH isomers content in animal-based foods according to NOVA classification. Red line represent median. Right: Kruskal-Wallis mean rank differences between groups at *p* = 0.05 significance level. **(D)** Left: dots plot showing 7-keto content in animal-based foods according to NOVA classification. Red line represent median. Right: Kruskal-Wallis mean rank differences between groups at *p* = 0.05 significance level. **(E)** Left: dots plot showing 5,6-epoxy isomers content in animal-based foods according to NOVA classification. Red line represent median. Right: Kruskal-Wallis mean rank differences between groups at *p* = 0.05 significance level. **(F)** Left: dots plot showing total phytosterols content in plant-based foods according to NOVA classification. Right: dots plot showing total DOxS content in plant-based foods for Groups 2 and 4 according to NOVA classification. Red line represent median.

### 3.3 DOxS content in animal and plant-based foods under WWEIA classification

This initial assessment demonstrates that, when large datasets are considered, ultra-processed foods contain a lower amount of DOxS compared to other NOVA groups. We, therefore, stratified animal-based foods according to the food source as per the WWEIA classification (**Fig. 4**). In WWEIA there are more than 160 unique categories assigned by a 4-digit number and description^32^. The Dietary Guidelines Advisory Committees (DGAC) has regrouped WWEIA categories in major categories according for analyses of contributions of food category intake to energy, nutrient and food group intakes^33^ (see Appendix E-2.7 in DGAC 2015 Advisory Report) (**Fig. 4A**). The FooDOxS database allows the stratification of DOxS data according to both grouping strategies. For this analysis, we focused on animal-based foods, since we have a higher number of reported data (**Fig. 2A**). Groups 6 (vegetables) and 3 (mixed dishes) showed the lowest amount of DOxS, compared to the high values observed for Groups 8 (condiments and dressings), Group 1 (dairy), as well as Group 7, which, for animal-based foods, consisted of protein-based nutritional beverages (category 7208 in the WWEIA classification) (**Fig. 4B**). Similar consideration can be made for 7-keto, a cholesterol derivative that has been considered a biomarker of food manufacturing, particularly thermal processes such as pasteurization and sterilization^24,34^. Group 8 (condiments and dressings) have significantly higher amount of 7-keto than Group 3 (mixed dishes) and Group 5 (snacks and sweets). It is worth noting that many ultra-processed foods fall into Groups 3 and 5 in the DGAC classification, which includes fast foods, ready-to-eat, restaurant items, and snacks. In summary, our findings suggest that, concerning DOxS accumulation, the influence of food processing appears to be less pronounced compared to the impact of food formulation. This implies that the specific composition and preparation of food play a more significant role in determining DOxS levels than the extent of processing.

**Figure 4.**
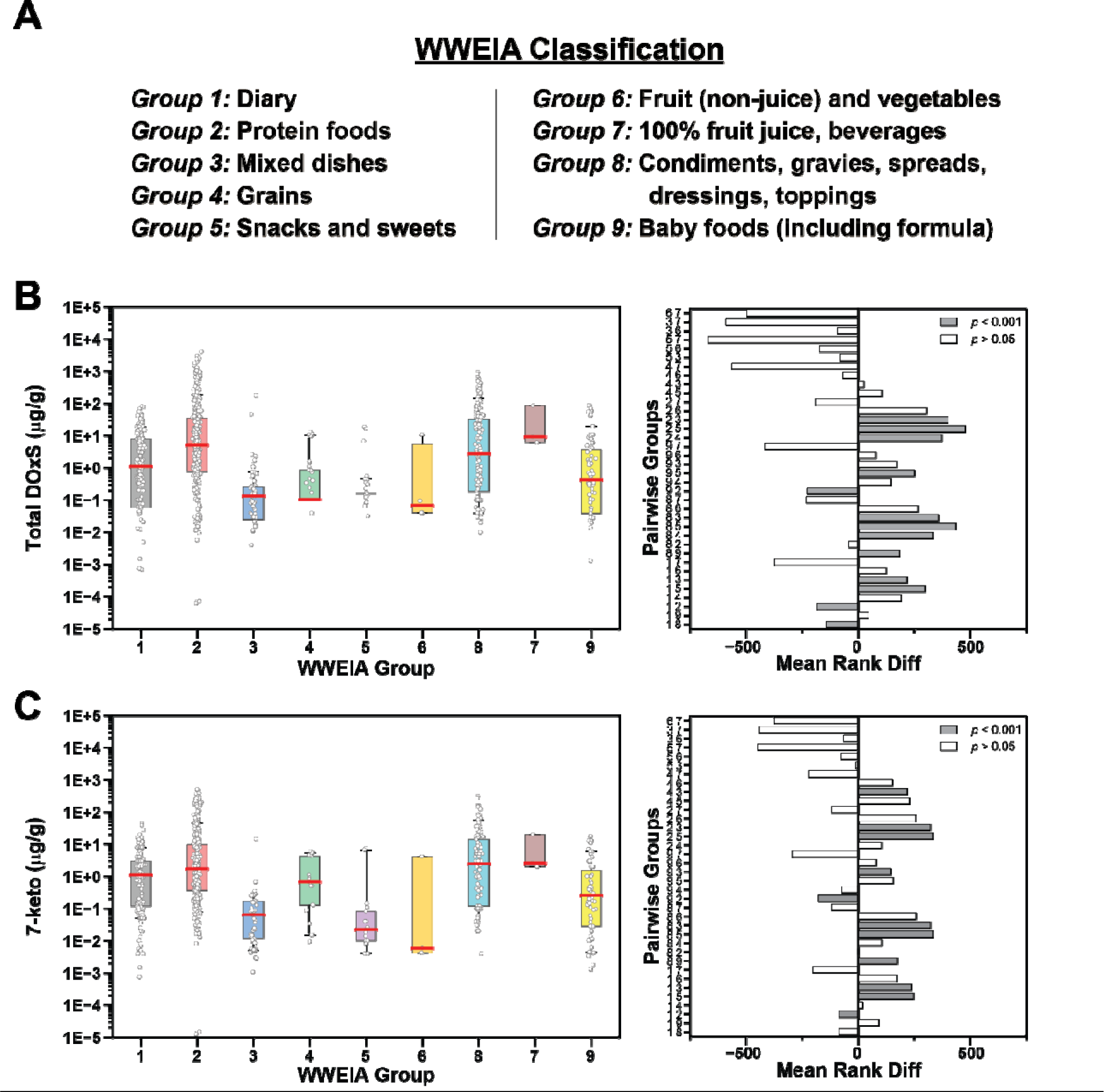
**DOxS abundance in FooDOxS database according to WWEIA classification**. **(A)** WWEIA macro-groups as simplified by the Dietary Guidelines Advisory Committees. **(B)** Left: Box plot showing total DOxS content in animal-based foods according to WWEIA classification. Box represents 25-75 confidence interval; red line represent median. Right: Kruskal-Wallis mean rank differences between groups at *p* = 0.05 significance level. **(C)** Left: Box plot showing 7-keto content in animal-based foods according to WWEIA classification. Box represents 25-75 confidence interval; red line represent median. Right: Kruskal-Wallis mean rank differences between groups at *p* = 0.05 significance level.

## DISCUSSION

### NOVA Classification and Criticisms

Industrial processing plays a pivotal role in ensuring both food safety and specific food characteristics. Processed foods have been integral to the human diet since the early stages of evolution, coinciding with the discovery of cooking, an original and fundamental processing technique^35^. However, in recent years, there has been a perceptible shift in consumer attitudes towards processed foods, with negative perceptions gaining traction^21^. This shift is influenced by several factors, including inadequate scientific understanding of food manufacturing, potentially misleading advertising, and recommendations from public health officials^21,36^. A defining moment in reshaping consumer awareness of food processing occurred in 2009 when Monteiro’s group in Brazil introduced the NOVA classification method^25^. This method, grounded in the degree of food processing, led to the identification of ’ultra-processed foods’, commonly abbreviated as UPFs. Over the years, various definitions for UPFs have emerged, with one of the latest emphasizing that these are formulations primarily composed of substances derived from foods and additives, with little to no intact Group 1 foods^26,37^. Monteiro’s group subsequently advocated for the adoption of NOVA by the UN Sustainable Development and its Decade of Nutrition initiative, using it as a guide for their food and health initiatives^20^.

The NOVA classification, particularly its characterization of UPFs, has faced criticism within the field of food science by prominent scientists^21,22,36,38^. Numerous critiques have been put forth regarding the classification criteria. Notably, products such as pasteurized milk, ultra-high temperature milk (UHT), and pasteurized juices are categorized as unprocessed or minimally processed (Group 1) foods according to NOVA. However, these items undergo high-energy-demanding processing conditions, including pasteurization and homogenization, challenging the notion of being unprocessed or minimally processed^36^. Regarding the definition of UPFs, one of the prominent criticisms lies in the association made by NOVA’s proponents between UPFs and low nutritional quality, leading to explicit recommendations to avoid their consumption^21^. This critique gains complexity when considering that UPFs encompass traditional foods like cheese and infant formulas, which are indispensable for infants not breastfed^36^.

### The FooDOxS database demystifies NOVA’s claims

The data presented in this paper challenges the prevailing notion that food processing, particularly within the framework of ’ultra-processing’ as defined by NOVA, has universally negative effects on food components. To scrutinize this hypothesis, we introduced FooDOxS, an extensive database encompassing oxidized sterols (DOxS) data for both animal- and plant-based foods. This is the largest database for DOxS in food products to date. The pivotal characteristic of DOxS lies in their predominantly processing-induced accumulation^9,23,24,34,39^. Various factors such as heat treatment, light exposure, and exposure to radical species derived from non-thermal processes, including light and non-thermal technologies, have the capacity to trigger the formation of DOxS^9,10,39^. Therefore, the accumulation of DOxS in foods serves as a distinctive signature of food processing, as extensively demonstrated by our group over the last decade. For instance, this is notably observed in infant formulations, where the spray-drying process to obtain powder formulas significantly enhances the occurrence of 7-ketocholesterol compared to liquid formulations^15,24^. Our database strongly demonstrates that, according to NOVA classification, animal-based ultra-processed foods contain less amounts of DOxS compared to all the other categories (**Fig. 3**). Particularly, UPFs contain less amounts of 7-ketocholesterol than unprocessed or minimally processed foods (**Fig. 3C**): this result is particularly interesting considering that 7-ketocholesterol is one of the most studied cholesterol-derived DOxS, with well-established bioactivity in humans^12,14,40^.

These findings prompt the question of whether the lower content of DOxS found in UPFs is a result of misattributed food items due to the superficial criteria of NOVA, as previously suggested^21,22^. To discern if DOxS content depends on the food source rather than processing, we stratified food items according to the WWEIA classification criteria, as simplified by the Dietary Guidelines Advisory Committees (DGAC) (**Fig. 4**). In WWEIA, processed foods are now distributed across several categories, with Group 3 (mixed dishes), Group 5 (sweets and snacks), and Group 8 (condiments and dressings) being the more representative. When grouped according to WWEIA, DOxS were more abundant in Group 2 (meat and seafood), Group 8 (condiments and dressings), and Group 1 (dairy), consistent with several reports obtained over several decades of DOxS research^9^. These results strongly suggest that the accumulation of sterols oxidative species is not strictly dependent on processing but rather on the formulation of the final food product.

### The FooDOxS database unveils gaps in DOxS data for specific food categories

We presented an extensive DOxS database featuring over 1,600 food items spanning various categories (**Fig. 2**). However, a notable discrepancy exists, with animal-based foods dominating, constituting over 75% of the total itemized foods (**Fig. 2A**). In contrast, plant-based food data are limited, primarily focusing on parental sterols, particularly phytosterols, with scant information on phytosterols-derived DOxS. This data gap is surprising given the ubiquity of vegetable oils rich in phytosterols in various foods, including infant formulas^15^. Multiple factors contribute to this data deficiency. Traditionally, cholesterol oxidation products have garnered more attention due to the prevalent belief in their significant role in human health and potential links to chronic diseases^9,39^. Additionally, the vast number of phytosterols, potentially yielding hundreds of oxidation products, poses challenges in their identification through analytical and spectroscopic methods^41^. Despite these challenges, the authors emphasize the urgency to address this data gap, especially considering the growing evidence of potential health effects associated with plant based DOxS, even at low concentrations^15–17,42^. The authors anticipate that the availability of pure standards for some of these oxidation derivatives will spur new data generation, leading to a more comprehensive understanding of their presence in foods.

## CONCLUSION

FooDOxS is an extensive dataset on oxidized sterols (DOxS) in both animal- and plant-based foods, compiled from literature and our laboratory. Using FooDOxS, we critically examine the NOVA classification, challenging the widely accepted notion that all processed foods, particularly ultra-processed foods (UPFs), universally have negative effects on food components. FooDOxS demonstrates that DOxS accumulation is more dependent on food formulation than the degree of processing, challenging NOVA’s claims, especially in its characterization of UPFs. Additionally, FooDOxS highlights a significant data gap in plant-based foods, with the majority of the over 1,600 presented food items being animal-based. This underscores the urgency for more comprehensive data on plant-based DOxS, especially considering the potential health effects associated with these compounds. In conclusion, FooDOxS serves as an open-access resource, offering valuable insights into the effects of food processing on DOxS content. Its implications extend to guiding food industry practices, informing policymakers, and aiding researchers in further understanding the exposure to DOxS in our diets.

## Data Availability

All data produced are available online at the FHEL GitHub webpage.

https://github.com/FHELMSU/FooDOxS

## Acknowledgements

The author would like to thank Lisa Zou, Ashley Xu, Lisaura Maldonado-Pereira, and Grant Gmitter for their valuable input. This study was partially funded by the Center for Research Ingredients Safety (CRIS) of Michigan State University with the GR100229 grant, and the USDA National Institute of Food and Agriculture, Hatch project MICL02526 to I.G.M.M. Y.V. was partially funded by the EnSURE program (2022 and 2023) from Michigan State University.

## Authors contribution

Y.V., Data curation, Visualization, Writing – Original Draft; C.B., Formal Analysis, Visualization, Writing – Original Draft, Writing – Review and Editing; I.G.M.M., Conceptualization, Project Administration, Resources, Supervision, Funding Acquisition, Writing – Review and Editing.

